# The Effects of Medication Strategies on Cardiovascular Outcomes in Patients with Marfan Syndrome: A Consistency Model Analysis

**DOI:** 10.1101/2023.03.27.23287828

**Authors:** Sih-Huan Huang, Chin-Yi Lin, Mei-Hwan Wu, Yi-No Kang, Hsin Hui-Chiu

## Abstract

**BACKGROUND:** Drug options for the treatment of Marfan syndrome (MFS) still warrant further investigation. Beta-blockers (BB) have long been considered the standard treatment, though several medications other than BB and angiotensin receptor blockers (ARB) have entered clinical practice and trials. However, no synthesis gathers evidence on medications other than BB or ARB. Therefore, this study aimed to investigate the effects and safety of medication strategies in managing MFS by synthesizing relevant randomized controlled trials (RCT).

**METHODS:** Three databases were searched for potential evidence using relevant keywords in both free-text and medical subject headings. Outcomes of interests were aortic root growth, aortic root Z score, aortic surgery, moderate-to-severe adverse events, cardiovascular and all-cause mortality. Quantitative data were pooled using frequentist-approach network meta-analysis in random-effects model.

**RESULTS:** Sixteen reports derived from 13 RCTs contributed to a seven-node consistency model including no treatment, BB, ARB, ARB+BB, calcium channel blockers (CCB), angiotensin converting enzyme inhibitor (ACEI), and combination of BB and renin inhibitor (RI). As compared with no treatment, RI+BB showed significant protection of aortic root growth (standardized mean difference [SMD]= -1.90, 95% confidence interval [CI]: -2.91, -0.89), followed by ARB+BB (SMD= -1.75, 95%CI: -2.40, -1.10). Nevertheless, no significant findings were seen in other clinical outcomes.

**CONCLUSION:** RI or ARB added on BB appear to be the optimal medication strategies to slow the progression of aortic root growth in MFS patients. However, we found no statistically significant difference in the risk of aortic surgeries, adverse effects, cardiovascular and all-cause mortality among medications. More RCTs with longer follow-up periods or bigger populations are needed to draw stronger evidence for clinical practices.

## INTRODUCTION

Marfan syndrome (MFS) is an inherited connective tissue disorder caused by fibrillin-1 (FBN1) gene mutation. Among all kinds of features, cardiovascular manifestations, especially progressive aortic root dilatation and subsequent aortic rupture or dissection, are the major concerns and the leading causes of death.^1^ Greater than 80% of Marfan patients have aortic root dilatation or mitral valve prolapse by the age of 18.^2^ Although aortic growth elevates risk of lethal and critical problems,^3^ the life expectancy of Marfan patients improved significantly due to the development of early diagnosis, medical treatment and prophylactic aortic surgery.^1, 4^ However, cardiovascular complications are still the noteworthy issues.^5, 6^

To date, beta-blockers (BB) remain the standard treatment for MFS and are recommended as early as possible after diagnosis due to amelioration of aortic dilatation.^7–11^ However, since activation of TGF-β signaling pathway was proved in fibrillin-1 deficient mouse models,^12–14^ various medicines, such as angiotensin receptor blockers (ARB), angiotensin converting enzyme inhibitor (ACEI), calcium channel blockers (CCB), renin inhibitors (RI), or combined therapy, have been widely investigated for MFS because of involvement in attenuating TGF-β signaling or other antihypertensive mechanisms.^15, 16^

Owing to the growing understanding of the pathophysiology of MFS, many different therapies have been widely discussed in these years, and recent syntheses on this topic have increased awareness of the use of ARB or combination of BB and ARB in the management of MFS. According to an important literature in the Lancet, there is some room for negotiation between the benefits of ARB alone and a combination of BB and ARB.^17^ Nevertheless, it appears to have no synthesis on the multi-treatment comparison of pharmacological managements (e.g. BB, ARB, ACEI, RI, and CCB) for MFS even though some relevant randomized controlled trials (RCTs) have been published in the past years. Therefore, the purpose of this study was to conduct a network meta-analysis to improve the understanding of which medication strategy for managing MFS is safe and the most effective as well.

## METHODS

This study was proposed and conducted according to the Preferred Reporting Items for Systematic Reviews and Meta-analysis (PRISMA) guidelines,^18^ and had been registered on the PROSPERO (CRD42022357777) before the completion of this study.

### Eligibility criteria and evidence selection

We reviewed RCTs from the literature that evaluated the outcomes of medical treatment in ameliorating risk of cardiovascular events in patients with MFS. The qualified trials for inclusion provided information concerning the following: (a) The inclusion and exclusion criteria for patient recruitment, (b) the regimen and dose of medical therapy, (c) the compression strategy used, (d) the measurement of aortic root growth, and (e) the evaluation of clinical outcomes. We excluded trials that met at least one of the following criteria: (a) The clinical outcomes had not been clearly stated; (b) the population was not Marfan syndrome; (c) the studied interventions were not medication therapies; or (d) the study design was not RCT.

Studies were identified by keyword searches of the Cochrane Library, Cochrane CENTRAL, EMBASE, and PubMed. The relevant terms were used in both free-text and medical subject headings (e.g. MeSH terms in PubMed and Emtree in EMBASE) including Marfan syndrome, angiotensin receptor blocker, angiotensin converting enzyme inhibitor, renin inhibitor, beta-blocker, as well as calcium channel blocker. Boolean operator “OR” and “AND” were applied to combine synonyms and identify intersection of two keywords set in terms of Marfan syndrome and medications respectively. The “related articles” facility in PubMed was used to broaden the search. No restriction of language or study types were applied. The last search was performed in October 2022. Appendix 1 detailed the database search strategy. Besides, additional studies by searching the reference sections of relevant papers and enquiring of known experts in the field.

Identified references were imported to a bibliographic software, Endnote X20 (Clarivate Analytics, US), in which duplicate of references were automatically routed out and the remainder were screened by two researchers (S.H.H. and C.Y.L.) based on the eligibility criteria. If they had any disagreements on the inclusions, an experienced researcher (Y.N.K) made the final decision through discussion after a research team meeting.

### Data extraction and quality evaluation

The two researchers (S.H.H. and C.Y.L.) independently extracted details of the RCTs pertaining to the sources of participants, inclusion and exclusion criteria, chosen prophylactic medication, arterial stiffness parameters, alteration in blood pressure, aortic root diameter growth, drug-related adverse effect, the incidence of aortic surgery, aortic dissection, cardiovascular death, and all-cause mortality. The individually recorded outcomes of the 2 reviewers were compared, and any disagreements were discussed and resolved based on the evaluation of an experienced researcher (Y.N.K).

The efficacy of pharmacological interventions was evaluated by continuous variables (pulse wave velocity, augmentation index, aortic root growth diameter, aortic root Z score), and dichotomous variables (the incidence of aortic surgery and dissection, adverse events, cardiovascular mortality, and all-cause mortality). Arterial stiffness was obtained by tonometry alone,^19, 20^ tonometry combined simultaneously with pulse wave Doppler and sphygmomanometer,^21^ or analyses of magnetic resonance imaging (MRI).^22^ According to the current American Society of Echocardiography guidelines,^23^ aortic parameters are usually measured at aortic valve annulus, the sinuses of Valsalva, and sinotubular junction. Among them, the maximal aortic diameter at the sinus of Valsalva, reported by the majority of the enrolled RCTs, was extracted for meta-analysis. Two different measuring methods were applied, cardiac echography or MRI. To adjust for somatic growth, aortic root Z score was calculated based on sinus of Valsalva diameter and body surface area.^24^ The number of patients underwent aortic surgery including prophylactic aortic root replacement and aortic dissection repair were documented.

The two researchers also independently appraised the methodological quality of each study based on the Revised Cochrane risk of bias tool for randomized trials (RoB 2) including the following domains: (a) randomization process, (b) deviations from the intended interventions, (c) missing outcome data, (d) measurement of the outcome, and (e) selection of the reported outcome.^25^ The final results were confirmed and agreed by research team meeting.

### Data synthesis and analysis

Some data of interests were tabulated if that information cannot be pooled statistically. The data were pooled only for studies that exhibited adequate clinical and methodological similarity, and quantitative synthesis was conducted using NMAStudio which has been developed as a web-based interactive tool for consistency model in the frequentist approach based on R package netmeta. Since some original RCTs did not report standard deviation of aortic root growth diameter and aortic root Z score,^11, 21, 26–28^ the present synthesis estimated the standard deviation based on the reported standard error, confidence interval (CI) limits, the first quartile, median, the third quartile, interquartile range, or range values. The effect sizes of the two synthesizable continuous outcomes were based on standardized mean difference (SMD) due to the variety in measuring aortic root growth diameter and Z score. For dichotomous data, when the incidence in both groups were zero, this synthesis imputed 0.5 for the sake of execution of analysis. The dichotomous outcomes were calculated using logarithm of risk ratio (RR), and back transformed to RR for result reporting. The precision of an effect size was reported as a 95% CI. Data was pooled in random-effects model no matter for SMD or RR. The clinically important size of effect was set to be 0.5 SMD. Statistical heterogeneity was assessed using the *I*^2^ statistics, with *I*^2^ quantifying the proportion of the total outcome variability that was attributable to variability among the studies. Incoherence of network model was tested using the Separate Indirect from Direct Evidence (SIDE) method, which is based on back-calculation.^29^ This synthesis also planned to detect small study effects using funnel plot and Egger’s regression intercept test when a pooled estimate was based on more than 10 studies. Certainty of evidence was further evaluated according to the methods of the Confidence In Network Meta-Analysis (CINeMA), and the confidence rating was only performed for those findings with statistical significance that may impact the clinical practice.

## RESULTS

Figure 1 displays the screening and selection process. We yield 1869 studies in initial search. Among all the studies, 385 studies were duplicated and 1449 were considered ineligible by screening the titles and abstract. In the remaining 35 references, 19 records were ruled out due to the following reasons: conference abstract without sufficient information and data (i = 4), not RCT (i = 12), and retraction (i = 3). The remained 16 eligible references were finally included in the present synthesis.^11, 19–22, 26–28, 30–37^

**Figure 1.**
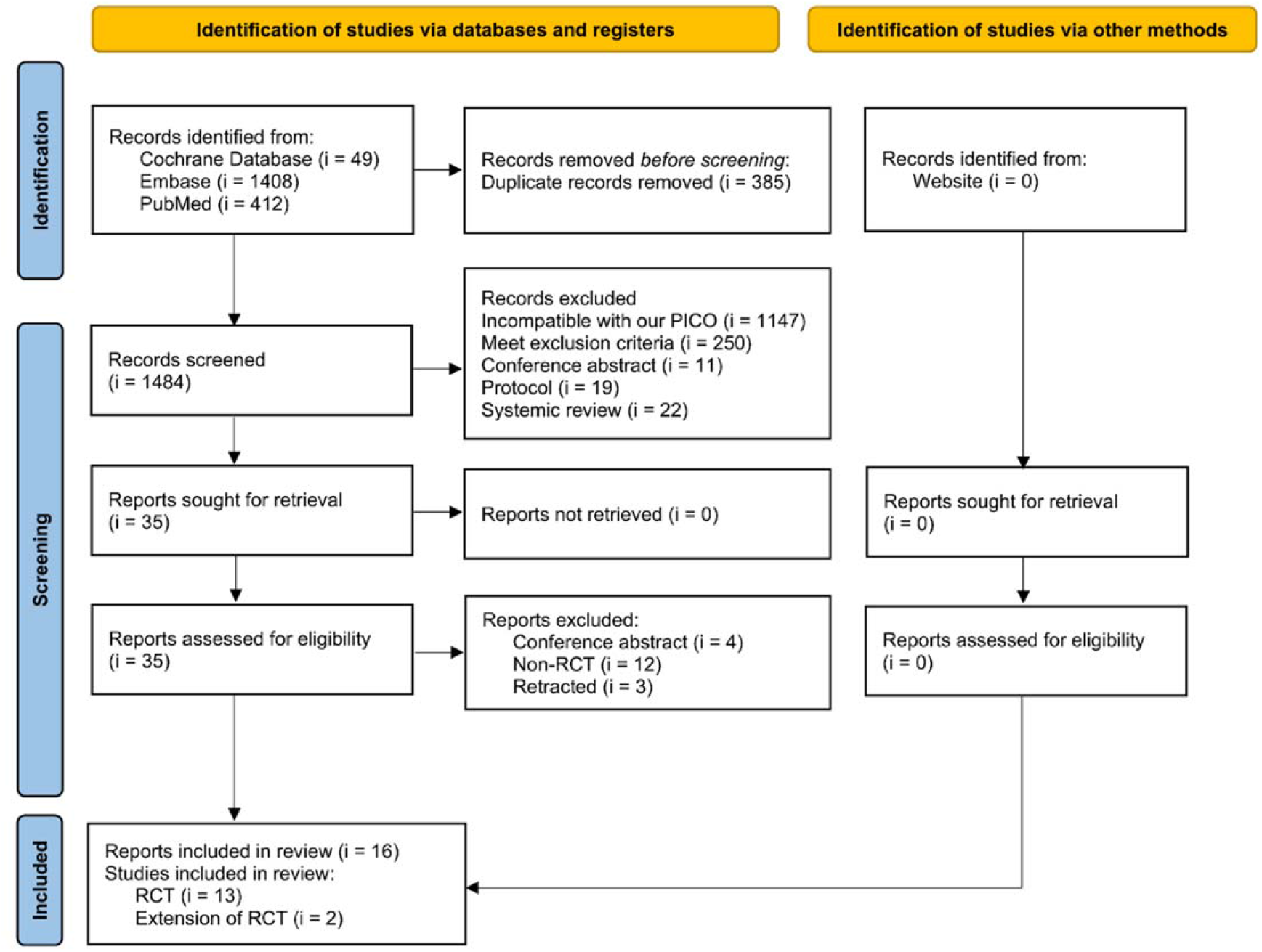
Flowchart of evidence selection for the synthesis of medications for managing Marfan syndrome. RCTs, randomized clinical trials.

### Characteristics and quality of included studies

The 16 references were based on 13 different RCTs and published in 1993 to 2020. Based on one of the pioneering clinical trials by Shore et al.,^11^ most RCTs implanted BB as baseline therapy in their control groups for ethical issue, and the trials contributed to a seven-node consistency model including no treatment, BB, ARB, ARB+BB, CCB, ACEI, and RI+BB. The characteristics of the RCTs were summarized in Table 1. Notably, although not included in the meta-analysis, two extended studies from established trials were also reviewed.^36, 37^

**Table 1.**
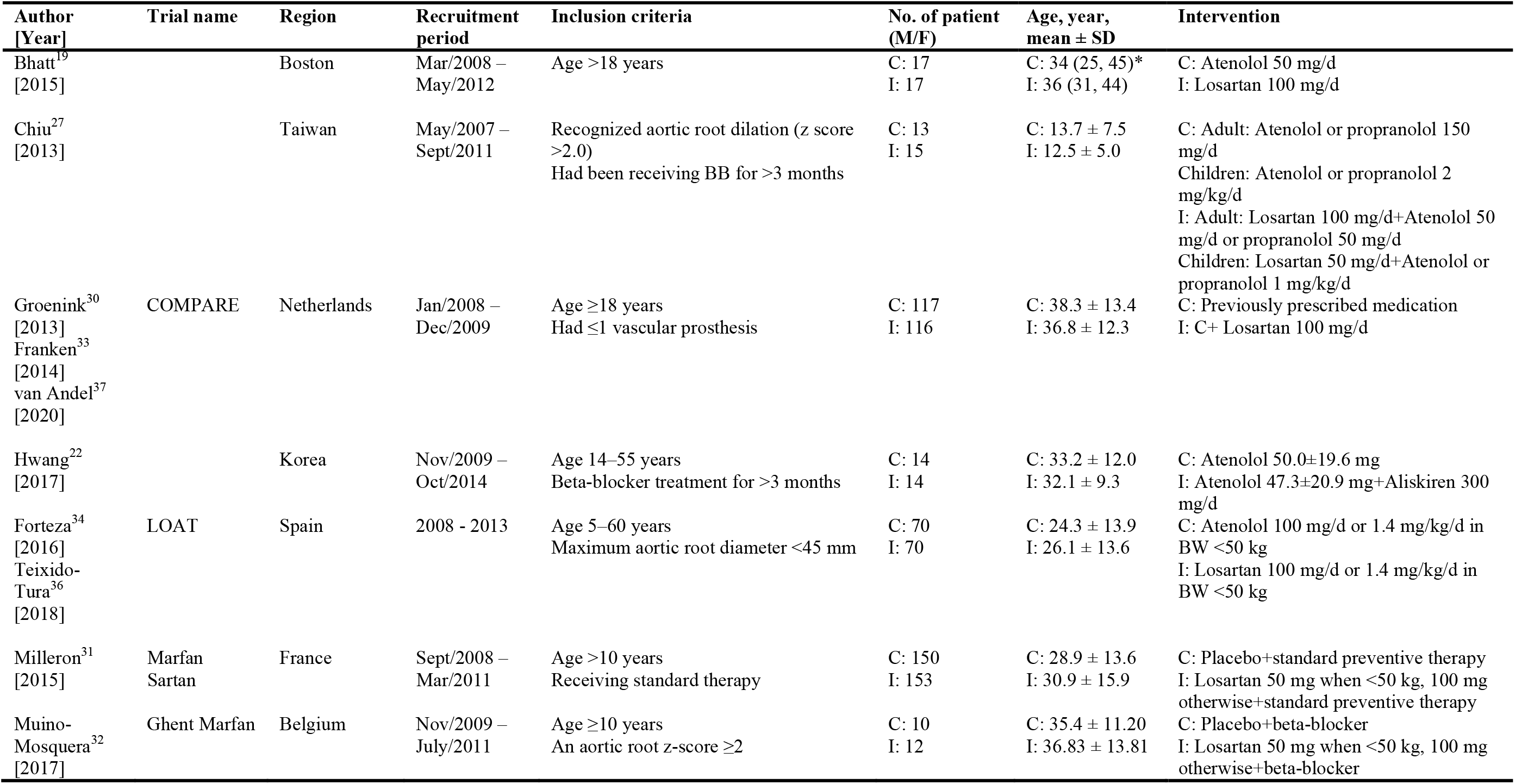
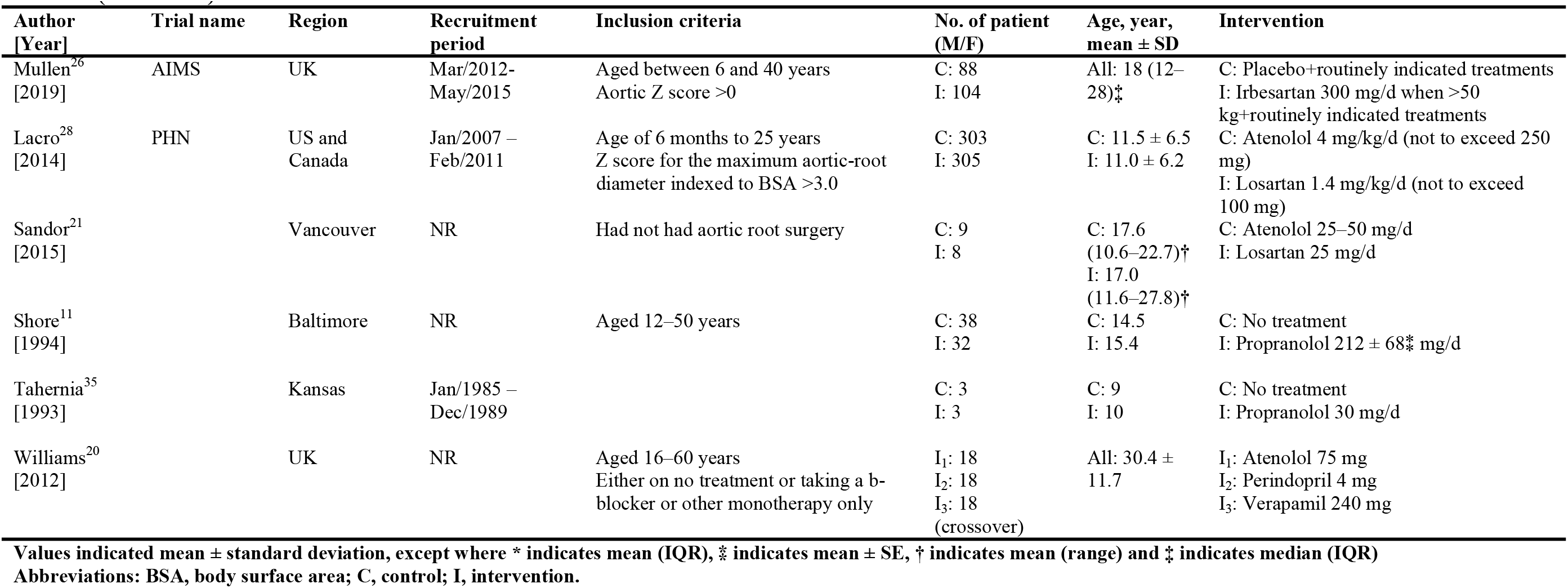
Characteristics of the included randomized controlled trials

**Table 2.** Summary of the findings of network evidence

| **Comparison** | **Aortic root**<br>**growth diameter**<br>**SMD (95% CI)** | **Aortic root**<br>**Z score**<br>**SMD (95% CI)** | **Aortic surgery**<br>**RR (95% CI)** | **Adverse event**<br>**RR (95% CI)** | **Cardiovascular**<br>**mortality**<br>**RR (95% CI)** | **All-cause**<br>**mortality**<br>**RR (95% CI)** |
| --- | --- | --- | --- | --- | --- | --- |
| RI+BB:no treatment | -1.90<br>(-2.91 to -0.89) |  |  | 0.41<br>(0.01 to 17.90) | 1.08<br>(0.01 to 114.78) | 0.43<br>(<0.01 to 38.02) |
| ACEI:no treatment | -1.60<br>(-2.60 to -0.59) |  |  | 2.04<br>(0.02 to 189.76) | 1.08<br>(0.01 to 116.29) | 0.43<br>(<0.01 to 38.54) |
| CCB:no treatment | -1.64<br>(-2.65 to -0.64) |  |  | 2.04<br>(0.02 to 189.76) | 1.08<br>(0.01 to 116.29) | 0.43<br>(<0.01 to 38.54) |
| ARB+BB:no treatment | -1.75<br>(-2.40 to -1.10) |  |  | 2.07<br>(0.19 to 22.54) | 0.96<br>(0.04 to 22.62) | 0.30<br>(0.02 to 4.75) |
| ARB:no treatment | -1.39<br>(-2.05 to -0.73) |  |  | 2.52<br>(0.23 to 27.81) | 1.08<br>(0.04 to 28.36) | 0.63<br>(0.03 to 11.77) |
| BB:no treatment | -1.45<br>(-2.06 to -0.84) |  |  | 2.04<br>(0.19 to 21.81) | 1.08<br>(0.08 to 15.07) | 0.43<br>(0.04 to 4.28) |
| RI+BB:BB | -0.44<br>(-1.25 to 0.36) | -0.29<br>(-1.03 to 0.45) |  | 0.20<br>(0.01 to 3.81) | 1.00<br>(0.02 to 46.96) | 1.00<br>(0.02 to 46.96) |
| ACEI:BB | -0.14<br>(-0.94 to 0.66) |  |  | 1.00<br>(0.02 to 47.71) | 1.00<br>(0.02 to 47.71) | 1.00<br>(0.02 to 47.71) |
| CCB:BB | -0.19<br>(-0.99 to 0.61) |  |  | 1.00<br>(0.02 to 47.71) | 1.00<br>(0.02 to 47.71) | 1.00<br>(0.02 to 47.71) |
| ARB+BB:BB | -0.30<br>(-0.53 to -0.07) | -0.12<br>(-0.30 to 0.06) | 1.03<br>(0.63 to 1.69) | 1.02<br>(0.78 to 1.33) | 0.89<br>(0.16 to 5.06) | 0.69<br>(0.15 to 3.26) |
| ARB:BB | 0.06<br>(-0.19 to 0.32) | -0.17<br>(-0.32 to -0.01) | 1.59<br>(0.77 to 3.27) | 1.24<br>(0.85 to 1.81) | 0.99<br>(0.14 to 6.93) | 1.46<br>(0.24 to 9.01) |

**Table 2 (continued) Summary of the findings of network evidence**
| Comparison | Aortic root<br>growth diameter<br>SMD (95% CI) | Aortic root<br>Z score<br>SMD (95% CI) | Aortic surgery<br>RR (95% CI) | Adverse event<br>RR (95% CI) | Cardiovascular<br>mortality<br>RR (95% CI) | All-cause<br>mortality<br>RR (95% CI) |
| --- | --- | --- | --- | --- | --- | --- |
| RI+BB:ARB | -0.51<br>(-1.35 to 0.34) | -0.12<br>(-0.88 to 0.64) | 0.65<br>(0.27 to 1.55) | 0.16<br>(0.01 to 3.16) | 1.01<br>(0.01 to 74.94) | 0.68<br>(0.01 to 48.29) |
| ACEI:ARB | -0.21<br>(-1.05 to 0.63) |  |  | 0.81<br>(0.02 to 39.31) | 1.01<br>(0.01 to 76.01) | 0.68<br>(0.01 to 48.99) |
| CCB:ARB | -0.26<br>(-1.09 to 0.58) |  |  | 0.81<br>(0.02 to 39.31) | 1.01<br>(0.01 to 76.01) | 0.68<br>(0.01 to 48.99) |
| ARB+BB:ARB | -0.36<br>(-0.70 to -0.02) | 0.05<br>(-0.19 to 0.28) |  | 0.82<br>(0.52 to 1.31) | 0.90<br>(0.07 to 12.12) | 0.47<br>(0.04 to 5.16) |
| RI+BB:ARB+BB | -0.14<br>(-0.98 to 0.70) | -0.17<br>(-0.94 to 0.60) |  | 0.20<br>(0.01 to 3.79) | 1.12<br>(0.02 to 76.52) | 1.45<br>(0.02 to 92.09) |
| ACEI:ARB+BB | 0.16<br>(-0.67 to 0.99) |  |  | 0.98<br>(0.02 to 47.33) | 1.12<br>(0.02 to 77.64) | 1.45<br>(0.02 to 93.45) |
| CCB:ARB+BB | 0.11<br>(-0.72 to 0.94) |  |  | 0.98<br>(0.02 to 47.33) | 1.12<br>(0.02 to 77.64) | 1.45<br>(0.02 to 93.45) |
| RI+BB:CCB | -0.25<br>(-1.39 to 0.88) |  |  | 0.20<br>(<0.01 to 25.83) | 1.00<br>(<0.01 to 233.90) | 1.00<br>(<0.01 to 233.90) |
| ACEI:CCB | 0.05<br>(-0.75 to 0.85) |  |  | 1.00<br>(0.02 to 47.71) | 1.00<br>(0.02 to 47.71) | 1.00<br>(0.02 to 47.71) |
| RI+BB:ACEI | -0.30<br>(-1.44 to 0.84) |  |  | 0.20<br>(<0.01 to 25.83) | 1.00<br>(<0.01 to 233.90) | 1.00<br>(<0.01 to 233.90) |

ACEI, angiotensin converting enzyme inhibitor; ARB, angiotensin receptor blocker; BB, beta-blocker; CCB, calcium channel blockers; CI, confidence interval; RI, renin inhibitor; RR, risk ratio; SMD, standardized mean difference.

**Table 3.**
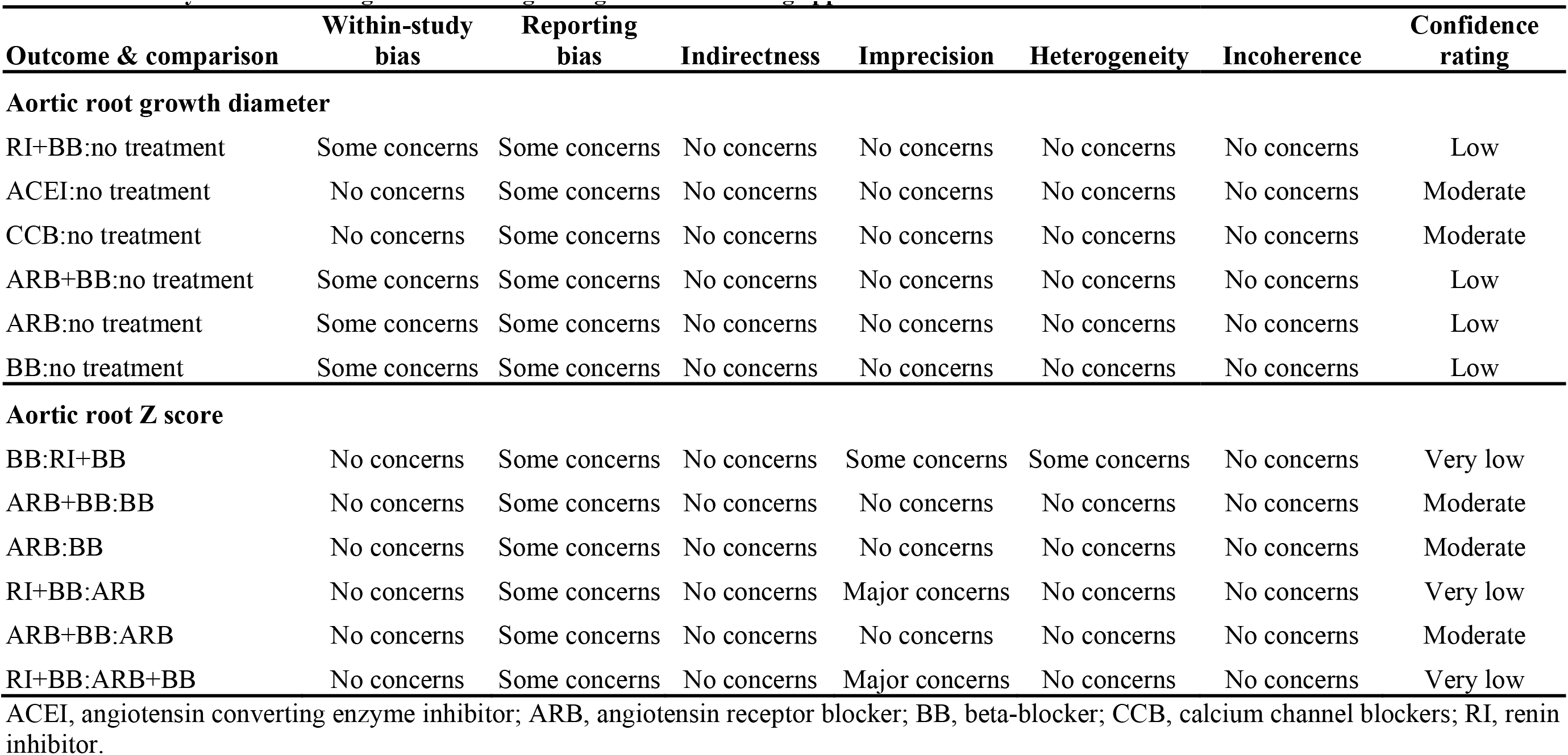
Certainty evaluation of significant findings using confidence rating approach

| **Outcome & comparison** | **Within-study bias** | **Reporting bias** | **Indirectness** | **Imprecision** | **Heterogeneity** | **Incoherence** | **Confidence rating** |
| --- | --- | --- | --- | --- | --- | --- | --- |
| **Aortic root growth diameter** |  |  |  |  |  |  |  |
| RI+BB:no treatment | Some concerns | Some concerns | No concerns | No concerns | No concerns | No concerns | Low |
| ACEI:no treatment | No concerns | Some concerns | No concerns | No concerns | No concerns | No concerns | Moderate |
| CCB:no treatment | No concerns | Some concerns | No concerns | No concerns | No concerns | No concerns | Moderate |
| ARB+BB:no treatment | Some concerns | Some concerns | No concerns | No concerns | No concerns | No concerns | Low |
| ARB:no treatment | Some concerns | Some concerns | No concerns | No concerns | No concerns | No concerns | Low |
| BB:no treatment | Some concerns | Some concerns | No concerns | No concerns | No concerns | No concerns | Low |
| **Aortic root Z score** |  |  |  |  |  |  |  |
| BB:RI+BB | No concerns | Some concerns | No concerns | Some concerns | Some concerns | No concerns | Very low |
| ARB+BB:BB | No concerns | Some concerns | No concerns | No concerns | No concerns | No concerns | Moderate |
| ARB:BB | No concerns | Some concerns | No concerns | No concerns | No concerns | No concerns | Moderate |
| RI+BB:ARB | No concerns | Some concerns | No concerns | Major concerns | No concerns | No concerns | Very low |
| ARB+BB:ARB | No concerns | Some concerns | No concerns | No concerns | No concerns | No concerns | Moderate |
| RI+BB:ARB+BB | No concerns | Some concerns | No concerns | Major concerns | No concerns | No concerns | Very low |

ACEI, angiotensin converting enzyme inhibitor; ARB, angiotensin receptor blocker; BB, beta-blocker; CCB, calcium channel blockers; RI, renin inhibitor.

Table S2 presents the methodological quality of included trials. Across all 13 trials, four of which have baseline imbalances suggesting a risk of bias in all the outcomes,^11, 21, 30, 32^ and two other trials did not explain the randomization process.^22, 35^ The number of patients who become lost to follow-up in extension of COMPARE study was greater than 20%.^37^ The interventions in Williams et al. administered for only four weeks,^20^ which we deem inappropriate to measure all-cause mortality and cardiovascular mortality. No obvious evidence to show serious bias due to selective result reporting in all 13 RCTs. Direct evidence of relevant outcomes have shown in Table S3 to S9.

### Rate of aortic dilatation

All 13 trials (n=1438) recorded the aortic dilatation rate or the aortic root diameter at the last follow up;^11, 19–22, 26–28, 30–32, 34–37^ however, we precluded the data from Tahernia et al. (n=6) due to negotiable definition with the wide variation of follow up period from 2 to 5 years.^35^ Besides, diversity was also noted in the assessment tool of aortic diameters; while most studies measured the aortic roots by echocardiography, the LOAT trial applied MRI only.^34^ Accordingly, a seven-node consistency model could be formed for rate of aortic root growth (Figure 2A). Regardless of the treatment regimen, the rate of aortic root growth was significantly lower in patients who received the medication (Table S10). In Figure 2B, it can be found that the aortic root growth rate was significantly reduced in RI+BB (SMD= -1.90; 95%CI: -2.91–-0.89), ARB+BB (SMD= -1.75; 95%CI: -2.40–-1.10), CCB (SMD= -1.64; 95%CI: -2.65–-0.64), ACEI (SMD= -1.60; 95%CI: -2.60–-0.59), BB alone (SMD= -1.45; 95%CI: -2.06–-0.84), and ARB alone (SMD= -1.39; 95%CI: -2.05–-0.73) as compared with no treatment. Combination of a renin-angiotensin system (RAS) inhibitor with BB seemed to have a more prominent effect in slowing aortic root dilatation than other single therapy when compared with no treatment. Similar trend of the effectiveness of the medication strategies was consistent with P-score (Figure 2C). However, there was no significant difference in the rate of aortic root growth amongst medication strategies.

**Figure 2.**
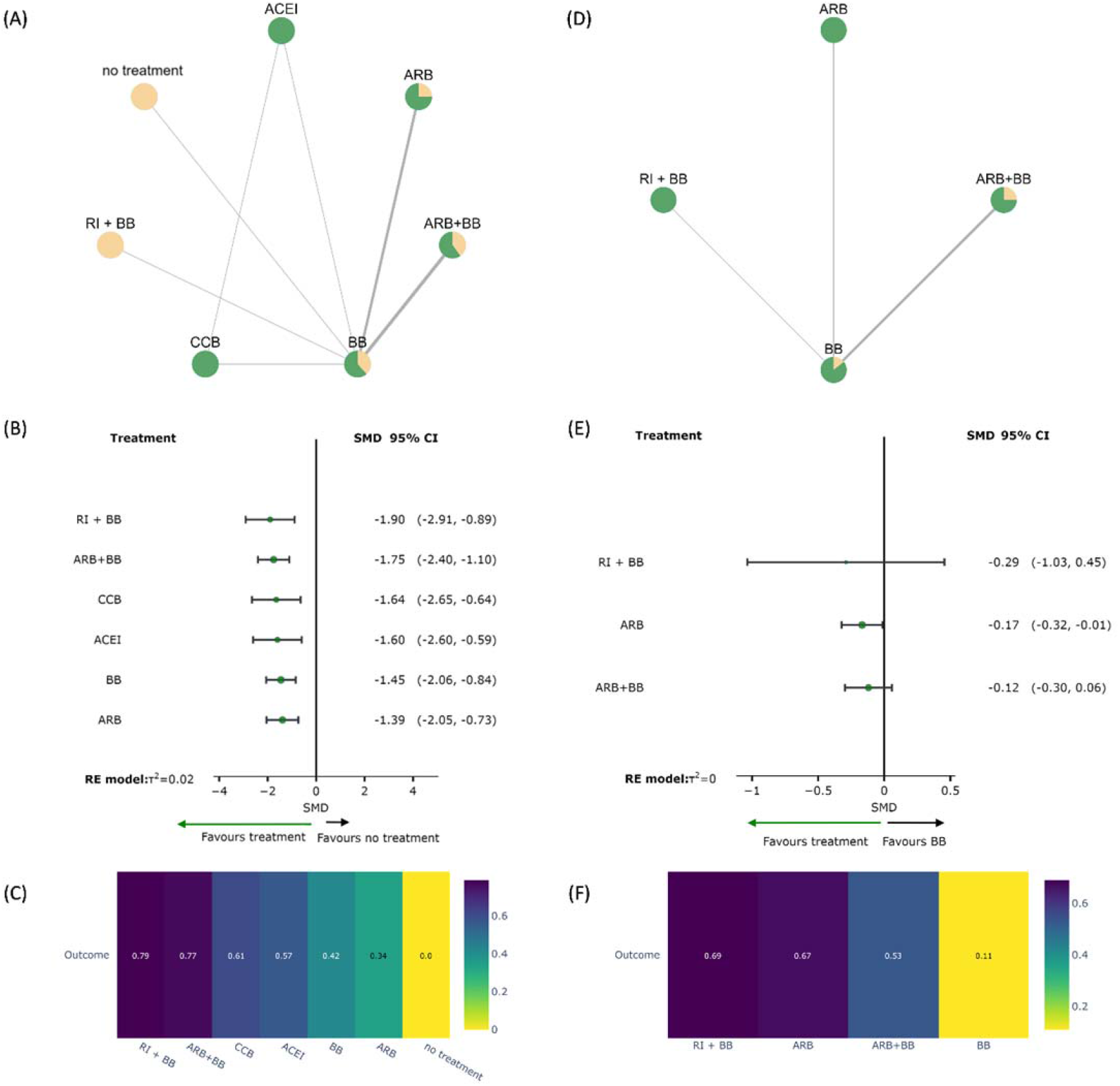
Network meta-analysis of aortic root diameter growth with (A) network plot, (B) forest plot, (C) P-score heat plot, and network meta-analysis of analysis of aortic root Z score with (D) network plot, (E) forest plot, and (F) P-score heat plot. ACEI, angiotensin converting enzyme inhibitor; ARB, angiotensin receptor blocker; BB, beta-blocker; CCB, calcium channel blockers; CI, confidence interval; RI, renin inhibitor; SMD= standardized mean difference.

Seven trials (n=1156) provided the data of aortic root Z score in their outcomes,^22, 26–28, 31, 32, 34^ and the data can form a four-node network but without closed loop (Figure 2D). Both dual regimens and ARB alone showed preferable outcomes over BB alone in alleviating the Z score change (Figure 2E). Worth noted, ARB alone (SMD= -0.17; 95%CI: -0.32–-0.01) performed significant benefit than BB alone. RI+BB was probably the best option, followed by ARB alone and ARB+BB (Figure 2F), although nonsignificant (Table S11).

### Aortic stiffness and left ventricle function

Parameters of arterial stiffness and left ventricle function were recorded in few studies and thus did not undergo meta-analysis. Remarkably, two RCTs,^19, 21^ enrolling 52 patients in total, comparing the impact of losartan and atenolol on vascular properties showed conflicting results. Pulse wave velocity (PWV) was significantly lowered by atenolol according to Bhatt et al.,^19^ while, despite a beneficial effect on hydraulic power, Sandor et al. noticed an increased PWV in the atenolol group.^21^ Decreased peripheral PWV via MRI was observed in aliskiren group compared with atenolol.^22^ However, no significant change in central PWV was demonstrated in patients treated with Perindopril or Verapamil.^20^ Augmentation index was found to be significantly decreased by losartan in one study.^19^ The disproportionate vascular load associated with increased aortic stiffness may contribute to left ventricular systolic and diastolic dysfunction, which can occur in adults with Marfan syndrome.^38^ Four studies measured left ventricular ejection fraction (LVEF),^19–21, 32^ only one showing significant better ejection fraction of atenolol group compared to losartan group at pre- and post-treatment.^21^ Most patients received 3-years of medical treatment and follow-up. The progression of aortic dilatation and clinical endpoints did not appear to differ between groups after long-term treatment.

### Clinical outcomes

Six trials (n=1494) analyzed the issue of aortic surgery,^26, 28, 30–32, 34^ most undergoing aortic root repair or replacement. Patients in two studies underwent elective or emergent surgery for aortic dissection,^26, 31^ and one patient in the study of Groenink et al. received prophylactic surgery of distal aorta.^30^ Nine trials monitored the efficacy of medication on aortic dissection.^11, 21, 26–28, 30–32, 34^ Although not significant, less patients underwent aortic surgery when treated with BB-alone. According to the 12 studies that provided mortality statistics,^11, 20–22, 26–28, 30–32, 34, 35^ none of the treatment regimens had a significant protective effect on cardiovascular or overall death. These 12 trials also reported the incidence of moderate to severe adverse events. Increased but non-significant RRs in adverse events were observed in the medication strategies. Table S12 to S16 showed results of consistency model for abovementioned clinical outcomes. No evidence showed serious small study effects and incoherence in all network analyses in this synthesis (Table S17 and S18).

## DISCUSSION

After full text review, fifteen RCTs with 1701 randomized patients were selected for analysis.^11, 19–22, 26–28, 30–32, 34–37^ Parameters in evaluating the efficacy of pharmacologic therapy in patients with MFS include aortic root diameter growth, standardized Z score, general adverse effect causing withdrawal, cardiovascular events (aortic dissection and aortic surgery), and mortality divided by cardiovascular etiology and all cause. In general, combined therapy with either ARB and BB or RI and BB outperform other monotherapies regarding each outcome, though without statistically significant difference. In continuous variables, RI exhibited greater advantage than ARB as add on regimen of BB. Conversely, ARB+BB was the most effective therapy in dichotomous variables. Remarkably, angiotensin with BB performed least effective in alleviating adverse events. None of the intervention showed statistically significant efficacy compared with the control group or BB except that monotherapy and combined therapy both are significantly more effective than no treatment in decreasing the rate of aortic root diameter growth.

Before our study, all meta-analyses looking into the medication control of Marfan syndrome lay focus on comparing the effect of ARB to BB. We reviewed the seven latest meta-analyses, and five of them concluded that the combination of ARB and BB most effectively slowing the aortic root dilatation (Table S19).^17, 39–42^ The inconsistency in the significance of the outcomes may be contributed to the different ways of classifying interventions. For instance, the experimental groups from three RCTs viewed as the ARB group by Malik et al. fell into the ARB and BB arm in our study. Similar difference can also be seen in the study by Wang et al., which could be possibly due to the confusion caused by ununified baseline therapy including BB, CCB, and others. Lack of significant difference in clinical outcomes is another feature shared by all. According to a Norwegian cohort study of 84 MFS patients recruited in 2003 and underwent new investigation from 2014 to 2015, the median cumulative probability of survival was 63 years for men and 73 years for women. Eleven of the 16 deaths were related to cardiovascular causes, among which eight were associated with aortic complications, including valve regurgitation due to dilatation. The median cumulative probability of aortic event-free survival in 63 patients included in the follow-up study was 37 years for men and 46 years for women.^43^ Likewise, population in Denmark and France demonstrated similar statistics, the former comprised 412 patients, in whom fifty percent were event free at age 49.6, median age at prophylactic surgery was 33.3 years and 41.2 years at dissection;^44^ after a mean follow-up period of 6.6 years, 732 patients from the latter studies bore an aortic risk of 0.10% per year when normalized aortic diameter < 20 mm/m2, 0.14% for 20 to 30 mm/m2, 0.43% for 30 to 42.5 mm/m2, and 5.07% for greater diameters.^45^ Due to the advanced age of aortic event free survival, the low annual aortic risk, and the guideline-derived recommendation of prophylactic aortic surgery once aortic root diameter ≥40-50 mm with risk factors,^46, 47^ all the RCTs included provided rare cases of aortic dissection and aortic surgery, leading to low statistical power and insignificant difference between groups. Moreover, prophylactic aortic surgery performed by clinicians’ discretion could mask the clinical outcomes in natural disease progression. Thirty-two percent and twenty-seven percent of the patients had previous aortic root replacement in three RCTs from two trials,^30, 32^ respectively, which may further enhance the heterogenicity of the enrolled studies.

### Pharmacological mechanisms of action in MFS patients

Aortic stiffness precedes and independently predicts thoracic aortic dilation in MFS, and implicates the risk of aortic dissection and rupture, which is associated with aortic diameter. Relevant mechanisms of RAS medications have been summarized in Figure 3, and we elaborate them in the following paragraphs. Carotid to femoral PWV relates directly to elastic modulus and is considered the gold standard of measuring arterial stiffness, also being the independent predicting factor of cardiovascular risk. Whereas augmentation index (Aix) is an indirect measurement of vascular stiffness, which is potentially influenced by factors that alter peripheral resistance, such as drugs or conditions.^48^ Bhatt et al. have observed significant effectiveness of atenolol in decreasing PWV and heart rate; losartan, in contrast, significantly lowering AIx.^19^ Although the precise mechanism via which BBs decrease PWV is unknown, reduction in blood pressure and transfer of stress from collagen to elastin may dampen stiffening of the aortic wall. Alternatively, the bradycardic effect of atenolol may also affect PWV by altering left ventricular ejection time. There was evidence that BB may protect the aortic root and reduce the risk of aortic dissection by reducing the impulse, namely, the rate of change in the central arterial pressure with respect to time (designated as dP/dt), of left ventricular ejection. In patients with malignant hypertension, methonium, which has a direct inotropic effect on the myocardium, increases impulse at a time when mean systemic pressure is low, leading to even more frequent occurrence of dissecting aneurysms than in untreated cases.^49^ Furthermore, several studies of animal model systems suggested that reducing the left ventricle impulse was much more protective than reducing mean blood pressure.^50^ Despite the negative chronotropic and inotropic actions of propranolol in benefiting the aorta, increased stiffness and decreased distensibility were observed in MFS patients with severely dilated aortic roots undergoing catheterization.^51, 52^ The possible explanations were different pharmacologic response in the high-risk patients from those with normal or moderately-dilated roots, and the requirement of long-term oral administration enabling the balance between reduction in the impulse, heart rate and elasticity over years. Ever since the two pioneering studies applicating propranolol in MFS patients,^11, 35^ the following RCTs alternated propranolol with atenolol, a β1-selective blocker with a longer half-life and fewer side effects, which may possess potential advantages over propranolol.

**Figure 3.**
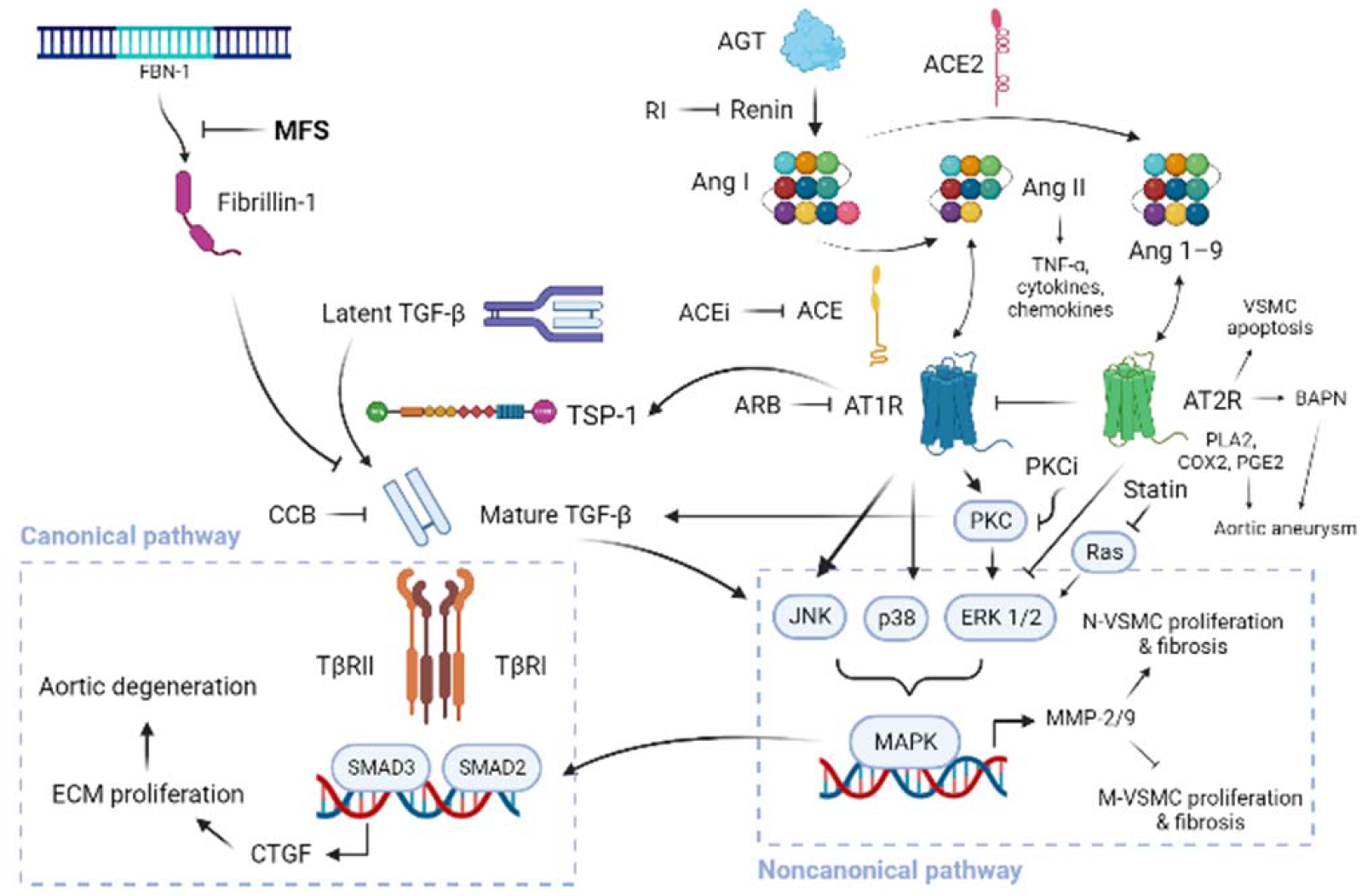
The pharmacological mechanism of action in MFS. The left part depicts the cascade of TGF-β signaling. Latent TGF-β is activated by TSP-1. The defect in FBN-1 gene in patients with MFS lead to the lack of fibrillin-1, which compounds with TGF-β and blocks the canonical pathway downstream causing aortic wall degeneration via SMAD2/3. The right-sided half shows the signal transferring from RAS to noncanonical pathway. At first, AT1R is considered to promote VSMC proliferation and fibrosis via MAPK, while AT2R, which inhibits AT1R, can preserve the function of VSMC. However, emerging evidences implies that AT2R may also transfer signals that cause VSMC apoptosis and increase the production of BAPN, which is correlated to aortic aneurysm formation. Additional medications, such as PKCi and statin, have also been proved to be effective in animal models. Created with BioRender.com. (ACE, angiotensin converting enzyme; AGT, angiotensinogen; Ang, angiotensin; AT1R, angiotensin I receptor; BAPN, β-aminopropionitrile monofumarate; COX, cyclooxygenase; CTGF, connective tissue growth factor; MMP, matrix metalloproteinase; M-VSMC, mesoderm-derived vascular smooth muscle cells; N-VSMC, neural crest-vascular smooth muscle cells; PGE, prostaglandin E; PKCi, protein kinase inhibitor; PLA, phospholipase A; TNF, tumor necrosis factor; TSP, thrombospondin)

As losartan did not change PWV in the study by Bhatt et al., it is inferred that reduced Aix was due to lowered peripheral impedance as a consequence of vascular smooth muscle relaxation.^19^ The property of losartan acting as an antagonist of TGF-β declared in mouse model of MFS is of greater importance.^53^ It has been well established that defective fibrillin-1 due to FBN-1 mutation in MFS fails to bind the latent complex of the cytokine TGF-β; excessive TGF-β signaling increases phosphorylation and nuclear translocation of downstream transcription factors Smad2/3,^54^ which regulate the production of connective tissue growth factor (CTGF) and collagens, resulting in elastic fiber fragmentation and aortic wall degeneration. TGF-β signaling cascades include both pathways (mitogen-activated protein kinase, predominantly extracellular signal-regulated kinase (ERK)1/2 but also c-Jun N-terminal protein kinase in some experimental contexts) by canonical (Smad-dependent) and by noncanonical (Smad-independent), two of them are activated in FBN-1 mutation mice in a TGFβ- and AT1 receptor–dependent manner.^55^ Angiotensin II binding to AT1 receptor can directly increases the expression of TGF-β ligands and receptors and indirectly induces the expression of thrombospondin-1, a potent activator of TGF-β.^56, 57^ In the vessel wall, AT1 signaling stimulates proliferation of vascular smooth muscle cells (VSMCs) and vessel wall fibrosis.^58^ While neural crest– and mesoderm-derived VSMCs (N- and M-VSMCs, respectively) respond conversely to TGF-β1 in avian systems, with cellular proliferation and fibrosis seen in the former and growth inhibition seen in the latter.^59^ This differential response may explain the particular predisposition of the aortic root—a vascular segment enriched for N-VSMCs—to undergo dilatation and dissection in MFS. Consistently, Groenink et al. observed significantly lower aortic root dilatation rate in the losartan group than in controls after 3.1±0.4 years of follow-up (0.77±1.36 vs. 1.35±1.55 mm, *P*=0.014), while no such benefit was generated in the trajectory beyond the aortic root.^30^

ACEI, which limits signaling through both AT1 and AT2 receptors, has been proven to be less protective than ARB in modifying MFS. Both drugs attenuated canonical TGF-β signaling in the aorta, but losartan uniquely inhibited TGFβ-mediated activation of ERK by allowing continued signaling through AT2, which correlated with the significantly higher therapeutic effects.^55^ However, whether AT2 contributes to aortic aneurysm development remains controversial in that AT2 signaling can both augment and inhibit the pathogenesis of aneurysm in preclinical models.^60^ Signaling through AT2R can oppose AT1-mediated enhancement of TGF-β signaling in some cell types and tissues,^61^ while it can also induce VSMC apoptosis, theoretically leading to aortic wall damage. Reduced Ang-II production with ACEI but selective AT1 receptor blockade ameliorated β-aminopropionitrile monofumarate-induced aortic aneurysm and dissection in rats, which is associated with increased expression of AT2 and VSMC apoptosis.^62^ Williams et al. noted a small but significant reduction in the diameter of the aorta at the level of the sinotubular junction after perindopril had been given for only 4 weeks.^20^ The interaction between AT1R and AT2R was further demonstrated by Habashi et al. with the use of ACEI in FBN1C1039G/+ mice or losartan in AT2KO: FBN1C1039G/+ mice, which results in no net change in ERK1/2 phosphorylation.^55^ This indicates that ongoing AT2R signaling is essential for the attenuation of ERK cascade on top of AT1R blockade, and that TGF-β-mediated ERK1/2 activation is the predominant driver of aneurysm progression in MFS mice. Thus, the ineffectiveness of enalapril on ERK reduction is probably attributable to the loss of AT2 receptor inhibitory signaling.^5^

CCB and ACEI reduce central systolic pressure and conduit arterial stiffness in adults with hypertension compared with BB.^63^ But similar effects in MFS have not been described. CCB, which have been proven to promote vascular remodeling and improve endothelial function, is considered an alternative if BB is intolerable, and may be more preferable due to its beneficial impact on vascular compliance.^64^ An estimation of 10% to 20% of MFS patients do not tolerate BB or are contraindicated because of asthma bronchiole, diabetes mellitus or depression.^5^ Verapamil is a non-dihydropyridine CCB with negative chronotropic and dromotropic properties, which reduces systolic pressure in the ascending aorta; and it also reduced AIx more, although non-significantly, than atenolol did.^20^ However, since CCB did not slow heart rate, it would not have any advantage over an ACEI in the treatment of patients with MFS. Beyond, the clinical evidence of safety and efficacy of CCB in MFS are still limited and under controversy. An in vitro study on TGF-β1-induced neonatal rat cardiac fibroblast showed combination treatment with R(−)efonidipine, an isomer of efonidipine that inhibits only the T-type calcium channel, and nifedipine, a selective L-type CCB, exerted complete attenuation of TGF-β1–induced collagen synthesis via Smad phosphorylation.^65^ While in a study of MFS mice, amlodipine and verapamil were both noticed to accelerate aortic aneurysm progression, dissection, and early mortality.^66^

Aliskiren is a direct RI and there is some evidence that aliskiren suppresses the expression and production of TGF-β in in vitro, in vivo and clinical studies.^67, 68^ Compared with ARB and ACEI, aliskiren has fewer adverse effects and may not lead to RAS “escape,” thus providing a greater RAS blockade. Another study has shown that co-administration of aliskiren and valsartan exerts a synergistic protection against renal fibrosis through the attenuation of messenger RNA expression of TGF-β in a unilateral ureteral obstruction rat model.^69^ Furthermore, RIs behave as vasodilators with the potential to improve the elasticity of the large arteries. Even though aliskiren did not improve aortic stiffness as measured by central PWV on MRI, brachial-ankle PWV was decreased following aliskiren treatment, which could be explained by the lowered central systolic blood pressure.^22^ Although FDA warned in 2012 about the possible risks of aliskiren in combination with ACEIs or ARBs in patients with diabetes or renal impairment, its use in other patients is relatively safe.^70^ By improving vascular stiffness via distinct mechanisms of action (Table S20), there is physiologic value to considering the use of combined medications.

### Drug-related side effects and combined use

Careful consideration and monitoring should be conducted when prescribing combined therapy to MFS patients. Two RCTs have reported cessation of losartan as add on treatment due to side effects like dizziness caused by low blood pressure, extreme fatigue, angioedema, or renal dysfunction.^30, 31^ Groenink et al. observed 17 out of 78 patients (22%) in losartan group prematurely discontinued the therapy, and targeted treatment dosage of 100 mg losartan daily was reached in only 54% of the patients, mainly due to side-effects, such as hypotension by concomitant BB use.^30^ According to the study by Milleron et al., six serious adverse events observed in four out of 151 patients receiving Losartan were considered to be potentially related to losartan. These events were pain related (lumbar, abdominal, and thoracic) or supraventricular tachycardia.^31^

BB, as a fundamental medication, do not target the pathogenic basis of aortic root dilation in MFS but simply aims to reduce hemodynamic stress on aortic vascular wall. Side effects of BB treatment such as fatigue, dizziness, headache, and constipation are a relevant problem during long-time treatment and lead to therapy cessation.^71^ Risk of inducing atrioventricular conduction delay by BB is another severe adverse effect requiring dose reduction as recorded by Shore et al.^11^ Much similarly, in a retrospective cohort study of 40 untreated pediatric patients with MFS adopted prophylactic ARB or BB treatment, BB was shifted to ARB in case of side effects at a mean age of 12.40±5.24 years in 22% patients; no patient had to stop ARB. While in four patients, BB therapy was supplemented by ARB in case of rapid progressive aortic root dilation at a mean age of 11.72±4.59 years (22%).^71^ Due to the different tolerance and susceptibility of side effects from person to person, the dose of BB prescribed varies greatly among individuals.

ARBs are used to treat hypertension and other cardiovascular diseases when ACEIs are not tolerated due to bradykinin-mediated side effects like dry cough and ACEI-induced angioedema, an uncommon but potentially serious side effect.^5, 72^ Diarrhea and nasopharyngitis were also reported in patients taking ramipril.^73^ Concern for adverse effects, including hyperkalemia, a rise in serum creatinine, and reduced glomerular filtration rate, indicates the need for regular monitoring under ACEI and ARBs use.^72^ Although ARBs do not inhibit bradykinin degradation, angioedema has been reported in association with their administration whether having experienced previous angioedema with an ACEI.^74, 75^ The more dominant side effects of aliskiren compared with ACEI were dizziness, headache, and cough.^73^ General symptoms associated with CCB use were dizziness, fatigue, and lightheadedness; children may present with non-specific signs such as vomiting, lethargy, hypotension. Non-dihydropyridines overdose usually presents with more severe symptoms, such as hypotension, bradycardia, jugular vein distention, altered level of consciousness, electrocardiogram changes, and worsening hyperglycemia. Clinicians should provide counseling on early signs of toxicity and medication safety for patients taking CCBs.^76^

Although there is limited research on the synergic effect of RAS inhibitor with BB, through their various mechanism of action, distinct adverse effects are prone to appear and change along with the duration of therapy. The optimal ways of drug selection, the priority and timing of their administration are still await being declared via more diverse designs of RCT study in the future.

### Limitation

With an aim to broaden the scope of medication being reviewed in Marfan patients, we enrolled two RCTs testing the efficacy of ACEI and RI, respectively. Nevertheless, due to the limited studies concerning drugs other than ARB, the results should be interpreted with caution. Another concern is the medication used as baseline therapy, although BB was the most frequently prescribed drug, CCB was also included.^30^ Specifically, patients taking BB accounted for only 54% in the irbesartan group and 59% in the placebo group.^26^ In other words, a small population classified into ARB+BB arm may not actually take the dual therapy. Since echocardiography may underestimated the maximum diameters in Marfan patients, who often bear asymmetric aortic roots, three enrolled RCTs applied MRI for aortic root growth measurement.^30, 32, 34^ Groenink et al. compared the two modalities and reported that while diameters measured by MRI were larger, aortic dilatation rate was comparable.^30^

## CONCLUSIONS

Our meta-analysis indicated that the addition of RI or ARB to BB appear to be optimal medication strategies for slowing the progression of aortic root growth in MFS patients. However, we found no statistically significant difference in the number of aortic surgeries, incidence of aortic dissection, adverse effects, cardiovascular and composite mortality in patients receiving single or dual medications. Overall, the methodological quality of the studies that we reviewed was good. Based on the results of our meta-analysis, more clinical trials concerning RI as add-on therapy to BB are anticipating to be performed in the future, possibly providing us with stronger evidence for clinical use.

## Data Availability

This is a network meta-analysis of randomized control trials published before October 31, 2022 from PubMed, EMBASE, Cochrane Library, and Cochrane CENTRAL.

## ACKNOWLEDGMENT

Financial support and sponsorship

This research waas supported in part by the National Science and Technology Council [MOST 108-2314-B-303 -020 –MY3] in Taiwan.

## Competing Interests

The authors declare that they have nothing to disclose regarding financial or non-financial conflicts of interest with respect to this manuscript.

## Provenance and peer review

Not commissioned, externally peer-reviewed.

## Notes

### Competing Interest Statement

The authors have declared no competing interest.

### Clinical Trial

PROSPERO (CRD42022357777)

### Funding Statement

This research was supported in part by the National Science and Technology Council [MOST 108-2314-B-303 -020 -MY3] in Taiwan.

### Author Declarations

This study was exempted from review by IRB of Taipei Tzu Chi Hospital.

